# Evaluation of Enzyme-Linked Immunoassay and Colloidal Gold-Immunochromatographic Assay Kit for Detection of Novel Coronavirus (SARS-Cov-2) Causing an Outbreak of Pneumonia (COVID-19)

**DOI:** 10.1101/2020.02.27.20028787

**Authors:** Jie Xiang, Mingzhe Yan, Hongze Li, Ting Liu, Chenyao Lin, Shuang Huang, Changxin Shen

## Abstract

**BACKGROUND:** In December 2019, a novel coronavirus (SARS-CoV-2)–infected pneumonia (COVID-19) occurred in Wuhan, China. Travel-associated cases have also been reported in other countries. The number of cases has increased rapidly but laboratory diagnosis is limited.

**METHODS:** We collect two groups of cases diagnosed with COVID-19 for experiments. One group collected 63 samples for Enzyme-linked immunosorbent assay (ELISA) IgG and IgM antibodies. The other group collected 91 plasma samples for colloidal gold-immunochromatographic assay (GICA).

**RESULTS:** The sensitivity of the combined ELISA IgM and ELISA IgG detection was 55/63 ( 87.3%), The sensitivity of the combined GICA IgM and GICA IgG detection was 75/91 ( 82.4%), Both methods are negative for healthy controls, specificity of 100%. There is no significant difference between the sensitivity of between ELISA and GICA (IgM+ IgG).

**CONCLUSIONS:** ELISA and GICA for specific IgM and IgG antibodies are conventional serological assays, they are simple, fast, and safe, the results can be used for clinical reference, and the huge clinical diagnosis and treatment pressure can be greatly relieved.

## Background

In December 2019, a cluster of acute respiratory illness, now known as novel coronavirus–infected pneumonia (NCIP), occurred in Wuhan, Hubei Province, China (1-3).The disease has rapidly spread from Wuhan to other areas. By February 27, 2020, there were 78630 confirmed cases reported in China, and tragically we have now surpassed 2747 people in China have lost their lives because of this virus. Most of the cases and most of the deaths are in Hubei province, Wuhan. Outside China, there are 3545 cases in 40 countries, and 35 death. The type of pneumonia caused by the 2019 novel coronavirus is a highly infectious disease, and the ongoing outbreak has been declared by WHO as a global public health emergency.

A novel coronavirus of zoonotic origin (2019-nCoV) has recently been identified in patients with acute respiratory disease. This virus is genetically similar to SARS coronavirus and bat SARS-like coronaviruses. Outbreaks in health care workers indicate human-to-human transmission. Full-genome sequencing and phylogenic analysis indicated that 2019-nCoV is a distinct clade from the beta coronaviruses associated with human severe acute respiratory syndrome (SARS) and Middle East respiratory syndrome (MERS)(1). International Committee on Taxonomy of Viruses(ICTV)is announced that 2019-nCoV is officially classified as severe acute respiratory syndrome coronavirus 2 (SARS-CoV-2)(4,5).The World Health Organization (WHO) announced that the official name of the disease caused by this virus is Corona Virus Disease (COVID-19)(6).

Huang et al first reported 41 cases of NCIP in which most patients had a history of exposure to Huanan Seafood Wholesale Market. Patients’ clinical manifestations included fever, nonproductive cough, dyspnea, myalgia, fatigue, normal or decreased leukocyte counts, and radiographic evidence of pneumonia. Organ dysfunction (eg, shock, acute respiratory distress syndrome (ARDS), acute cardiac injury, and acute kidney injury and death can occur in severe cases(2). Although most patients present with mild febrile illness with patchy pulmonary inflammation, a significant portion develop severe ARDS, with a current case fatality of 4.3%(3).

Diagnosis is based on clinical history and laboratory and chest radiographic findings, but confirmation currently relies on nucleic acid-based assays. The nucleic acid-based assay has been used to confirm the diagnosis of new pneumonia, but many patients will miss the diagnosis. We have verified the two new reagents kit that are currently not on the market and have achieved good results. The following are reported as follows:

## Materials and Methods

### Study Design and Participants

This case series was approved by the institutional ethics board of Wuhan Jinyintan Hospital. All consecutive patients with confirmed COVID-19 admitted to Jinyintan Hospital from January 1 to January 28, 2020, were enrolled. Oral consent was obtained from patients. Jinyintan Hospital, located in Wuhan, Hubei Province, the endemic areas of COVID-19, is first designated hospital for COVID-19 patients in Wuhan. All patients with COVID-19 enrolled in this study were diagnosed according to World Health Organization interim guidance (7, 8). The remaining serum samples for clinical testing were collected from two groups of patients. One group collected 63 samples from February 2, 2020 for Enzyme-linked immunosorbent assay (ELISA) IgG and IgM antibodies. The other group collected 91 plasma samples from February 3-4, 2020 for colloidal gold-immunochromatographic assay (GICA), of which 81 cases took throat swab sample for nucleic acids (Real-Time Reverse Transcription Polymerase Chain Reaction Assay, qRT-PCR) detection. 35 healthy individuals served as controls in both groups.

### ELISA

The novel coronavirus IgG/IgM antibody ELISA kits (lot number 2020010108) were manufactured by Zhu Hai Liv Zon Diagnostics Inc. The 98 serum samples were tested according to the manufacturer’s instructions. IgM capture ELISA: Firstly, 5 μL serum samples were diluted in 500 μL sample diluent. Then added 100 μL diluted samples to duplicate wells of microplates which were coated with mouse anti-human IgM monoclonal antibody (μchain), and incubated for 60 min at 37□±1□. Plates were washed five times and reacted with 100 μL enzyme marker (enzyme-labeled antibody-linked antigen) for 30 min at 37□±1°C for the purpose of detecting IgM against new coronavirus in serum samples. IgG indirect ELISA:5 μL serum samples diluted in 100 μL sample diluent were added to duplicate wells of microplates which were coated with recombinant antigen of new coronavirus, and incubated for 60 min at 37□±1□. Plates were washed five times and reacted with 100 μL enzyme marker (HRP-conjugated monoclonal mouse anti-human IgG) for 30 min at 37□±1 °C for the purpose of detecting IgG against new coronavirus in serum samples. Plates were then washed five times, and 50 μL substrate buffer and 50 μL tetramethylbenzidine (TMB) substrate solution were added to each well for a chromogenic reaction for 15 min at 37□±1 °C. The color reaction was stopped by the addition 50 μL of 2 M H2SO4 to each well. Finally, the OD450 was measured and recorded immediately using an Infinite 200 PRO microplatereader.

### GICA

The novel coronavirus IgG/IgM antibody GICA kits (lot number 2001010220) were manufactured by Zhu Hai Liv Zon Diagnostics Inc, China. The 126 serum samples were tested according to the manufacturer’s instructions. For each test, 10μL of serum sample and 100μL of sample diluent were added vertically onto the sample pad of the test strip. The strip was then placed flat to allow the solution to migrate up the membrane, through gold-labeled pad, the testing area, quality control area and finally to adsorption zone.

After 10 min, the result was judged by the color of the test and control lines. If both the detection band and the control band turned red, the sample was recorded as positive. If the control band turned red but the detection band was not colored, it was recorded as negative. If neither band was colored, the test reagents were assumed to be not working.

### qRT-PCR Assay for SARS-CoV-2

Throat swab samples, sputum samples and alveolar lavage fluid samples were collected for extracting 2019-nCoV RNA from patients suspected of having 2019-nCoV infection. After collection, the throat swabs were placed into a sterile test tube with 1 ml sterile saline, the sputum samples were added equal volume of acetylcysteine (10g/L) and shaken at room temperature for 30 min to be fully liquefied, and total RNA was extracted using the nucleic acid extraction kit (QIAamp viral RNA mini kit). In brief, 40μL of cell lysates were transferred into a collection tube followed by vortex for 10 seconds. After standing at room temperature for 10 minutes, the collection tube was centrifugated at 1000 rpm/min for 5 minutes. The suspension was used for qRT-PCR assay of 2019-nCoV RNA. Then, n*19 μL mixed reagent of fluorescence PCR detection for 2019-nCoV nucleic acid and n* 1 μL RT-PCR enzyme (n was the number of reaction tubes) were mixed and vortexed for a few seconds. The above mixture of 20 μL was put into the PCR reaction tube respectively, after that the extracted sample by 5 μL was added. qRT-PCR analysis was conducted using the ABI 7500 Real-Time PCR System. The PCR parameters were 45 °C for 10 min, 95 °C for 3 min, followed by 45 cycles of 95 °C for 15 s, 58 °C for 30 s, and a single fluorescence detection point at 58°C. Two target genes, including open reading frame 1ab (ORF1ab) and nucleocapsid protein (N), were simultaneously amplified and tested during the qRT-PCR assay. The qRT-PCR assay was performed using a 2019-nCoV nucleic acid detection kit according to the manufacturer’s protocol (Shanghai ZJ Bio-Tech Co Ltd). A cycle threshold value (Ct-value) less than 37 was defined as a positive test result, and a Ct-value of 40 or more was defined as a negative test. These diagnostic criteria were based on the recommendation by the National Institute for Viral Disease Control and Prevention (China) (http://ivdc.chinacdc.cn/kyjz/202001/t20200121_211337.html). A medium load, defined as a Ct-value of 37 to less than 40, required confirmation by retesting.

### Statistical Analysis

All analyses were performed using SPSS 22.0. Categorical variables were expressed as frequencies (percentages), and performed using the chi-square test with Yates’s correction or Fisher’s exact test, as appropriate. P<0.05 was considered statistically significant. For multiple statistical comparisons, chi-square test was corrected by Bonferroni’s correction (0.05/test numbers).

## Results

### ELISA results

ELISA test of 63 samples of known COVID-19 patients, the results showed that 28 IgM antibodies were positive, the sensitivity was 44.4% (28/63), the specificity was 100% (35/35), the accuracy was 63/98 (64.3). 52 IgG antibodies were positive, with an accuracy of 82.54% (52/63), specificity of 100% (35/35), and accuracy of 87/98 (88.8). The sensitivity of the combined IgM and IgG detection was 55/63 (87.3%) (Table 1). (Supplementary Table 1 and Table 2)

**Table 1.** Sensitivity, specificity and accuracy of ELISA IgM, IgG,when tested independently or in combinations

| Methods | Sensitivity (%) | Specificity (%) | Accuracy (%) |
| --- | --- | --- | --- |
| ELISA IgM | 28/63(44.4) | 35/35 ( 100.0) | 63/98(64.3) |
| ELISA IgG | 52/63(82.5) | 35/35 ( 100.0) | 87/98(88.8) |
| ELISA IgM+ IgG* | 55/63(87.3) | 35/35 ( 100.0) | 90/98(91.8) |

\* positive if any of two markers is positive

### GICA results

GICA test of 91 samples of known COVID-19 patients, the results showed that 52 IgM antibodies were positive, the sensitivity was 57.1% (52/91), the specificity was 100% (35/35), the accuracy was 69.0% (87/126). 74 IgG antibodies were positive, with an accuracy of 81.3% (74/91), specificity of 100% (35/35), and accuracy of 109/126 (86.5). The sensitivity of the combined IgM and IgG detection was 75/91 (82.4%) (Table 2). (Supplementary Table 3 and Table 4)

**Table 2.** Sensitivity, specificity and accuracy of colloidal gold IgM, IgG,when tested independently or in combinations

| Methods | Sensitivity (%) | Specificity (%) | Accuracy (%) |
| --- | --- | --- | --- |
| colloidal gold IgM | 52/91(57.1) | 35/35 ( 100.0) | 87/126(69.0) |
| colloidal gold IgG | 74/91(81.3) | 35/35 ( 100.0) | 109/126(86.5) |
| colloidal gold IgM+ IgG* | 75/91(82.4) | 35/35 ( 100.0) | 110/126(87.3) |

\* positive if any of two markers is positive

### qRT-PCR results

qRT-PCR test of 81 samples of known COVID-19 patients, the results showed that 42 cases were positive, the sensitivity was 51.9% (42/81).

### Compare the sensitivity of the three methods (ELISA IgM+ IgG, GICA IgM+IgG, qRT-PCR)

Overall, there are significant differences in the three detection methods (P<0.001). There is no significant difference between the sensitivity of between ELISA (IgM+ IgG) and GICA (IgM+ IgG), P=0.411. (Table 3)

**Table 3.** Compare the sensitivity of the three methods

| Methods | Postivie | Negative | Total | P |
| --- | --- | --- | --- | --- |
| ELISA IgM+ IgG | 55 ( 87.3% ) | 8 ( 12.7% ) | 63 | <0.001<sup>*</sup> |
| GICA IgM+ IgG | 75 ( 82.4% ) | 16 ( 17.6% ) | 91 | 0.411<sup>**</sup> |
| qRT-PCR | 42 ( 51.9% ) | 39 ( 48.1% ) | 81 |  |

<sup>\*</sup>P<0.001 , Overall, there are significant differences in the three detection methods.
<sup>\*\*</sup>P=0.411 , No significant difference between ELISA (IgM+ IgG) and GICA (IgM+ IgG)

## Discussion

At present, the sample collection for nucleic acid detection of suspected cases of COVID-19 is mostly upper respiratory tract samples (mainly throat swabs) (9). The collection of throat swabs is not standardized and it is easy to miss the diagnosis. The collection process is extremely risky for medical staff. Confirmed diagnosis is playing an important role in facilitating patient isolation, treatment and assessment of infectious activities. However, due to their limited capacity to handle an epidemic of the current scale and insufficient supply of assay kits, only a portion of suspected cases can be tested, leading to incompleteness and inaccuracy in updating new cases, as well as delayed diagnosis. A fast-performing serologic assay is acutely needed for the current and outbreak.

In this study, we tested serum samples from confirmed COVID-19 patients with two assay kits and achieved a high positive rate. The sensitivity of the combined ELISA IgM and IgG detection was 55/63 (87.3%), The sensitivity of the combined GICA IgM and IgG detection was 75/91 (82.4%), and the healthy controls were negative. The case group was COVID-19 patients diagnosed by qRT-PCR. Since many patients are undergoing treatment, they are gradually recovering, the nucleic acid test result has become negative, so the positive rate of patients tested was only 51.9%.

It is worth noting that the new type of coronavirus antibody of the kit is against the severe acute respiratory syndrome (SARS)-like coronavirus, not only against SARS-CoV-2, but the IgG originally infected with SARS may also be positive. But our research found, the healthy controls are all negative and the specificity is very good.

ELISA and GICA for specific IgM and IgG antibodies is conventional serological assays, they can offer a high-throughput alternative, which allows for uniform tests for all suspected patients, and can facilitate more complete identification of infected cases and avoidance of unnecessary cross infection among unselected patients. They use plasma or serum as test samples, and blood samples can be collected easily, which can greatly reduce the risk of infection for medical staff. The complicated processing procedure of the sample during the laboratory test is removed, the test result can be obtained quickly, the operation is simple, the safety of medical staff can be protected, and the huge clinical diagnosis and treatment pressure can be greatly relieved.

Although ELISA and GICA are simple, fast, and safe, the results can be used for clinical reference. Cases confirmation still depended on qRT-PCR, and analyze epidemiological, demographic, clinical, and radiological features and other laboratory data. There is a need to further increase the number of samples tested to prove their t of sample testing to confirm their significance.

In conclusion, ELISA and GICA for specific IgM and IgG antibodies is conventional serological assays, they are simple, fast, and safe for diagnosis COVID-19. The results can be used for clinical reference, and the huge clinical diagnosis and treatment pressure can be greatly relieved.

## Data Availability

All data generated or analyzed during this study are included in this published article and its supplementary information files.

## Acknowledgements

This work was supported by the Zhongnan Hospital of Wuhan University Science, Technology and Innovation Seed Fund under Grant znpy2017022.

